# Self-Reported Fitness (IFIS) in Adults with Type 2 Diabetes Mellitus: Validity and Associations with Cardiometabolic and Body Composition Outcomes

**DOI:** 10.1101/2025.01.17.25320713

**Authors:** Ángel Herraiz-Adillo, Karin Rådholm, Fredrik Iredahl, Carl-Johan Carlhäll, Nahuel Roemkens, Mikael Forsgren, Olof Dahlqvist Leinhard, Peter Lundberg, Stergios Kechagias, Nils Dahlström, Patrik Nasr, Mattias Ekstedt, Pontus Henriksson

## Abstract

This study aimed to assess the validity of the International Fitness Scale (IFIS) for evaluating cardiorespiratory fitness compared to the 6-minute walk test (criterion validity) and to examine their associations with cardiometabolic and body composition outcomes (construct validity) in adults with type 2 diabetes mellitus (T2DM). A cross-sectional analysis was conducted on 282 adults with T2DM (mean age 63.6 ± 8.1 years, 37.9% women). Self-reported fitness was assessed using IFIS, including overall fitness and cardiorespiratory fitness scores. Objective cardiorespiratory fitness was assessed using the 6-minute walk test. Associations with cardiometabolic (i.e., cardiovascular health score, metabolic dysfunction-associated steatotic liver disease [MASLD], high-sensitive C-reactive protein) and body composition outcomes (i.e., body mass index [BMI], visceral adipose tissue volume, and thigh fat-free muscle volume) were analyzed using ANCOVA and ROC curves, which were compared using DeLong tests.

Both IFIS overall and cardiorespiratory fitness strongly correlated with performance in the 6-minute walk test (p<0.001 for ANCOVA models) and were associated with all cardiometabolic and body composition outcomes. ROC analyses showed at least similar predictive performance for IFIS overall and cardiorespiratory fitness scores compared to the 6-minute walk test. Furthermore, IFIS overall outperformed the 6-minute walk test for predicting poor cardiovascular health (AUCs: 0.678 [95% CI: 0.618-0.737] versus 0.586 [95% CI: 0.517-0.654]) as well as MASLD (AUCs: 0.630 [95% CI: 0.563-0.696] versus 0.519 [95% CI: 0.443-0.595]).

This study proves that IFIS is a valid tool for assessing physical fitness in T2DM patients and that it conveys cardiometabolic and body composition outcomes at least as well as the 6-minute walk test.

## INTRODUCTION

Type 2 diabetes mellitus (T2DM) is a growing global health concern, with its prevalence rising alarmingly^1^ and affecting more than 500 million people.^2^ While the etiology of T2DM is multifactorial, it is closely associated with lifestyle factors including obesity, poor diet, and physical inactivity. Individuals with T2DM are at a significantly higher risk of developing cardiovascular risk factors (e.g., hypertension, dyslipidemia, insulin resistance, and liver steatosis) and cardiovascular disease (CVD).^3^

Adequate physical fitness, particularly cardiorespiratory fitness, has been associated with a reduced risk of developing T2DM and with improved outcomes in patients already diagnosed with the condition.^4,5^ The American Heart Association recognizes cardiorespiratory fitness as a vital clinical sign and advocates for its routine assessment in clinical settings.^5^ However, laboratory cardiopulmonary exercise testing, the gold standard for evaluating cardiorespiratory fitness, is often impractical in routine clinical practice due to substantial resource demands related to personnel, equipment, and costs. While some field exercise tests, including the 20-meter and the 6-minute walk tests,^6^ can objectively assess cardiorespiratory fitness, they may be impractical in clinical practice, particularly for patients with T2DM. These patients may experience conditions like peripheral neuropathy or peripheral arterial disease, which make walking tests challenging. This highlights the need for more accessible and less resource-intensive methods, such as self-reported cardiorespiratory fitness assessments.

Previous studies have demonstrated that self-reported physical fitness, particularly through the use of the International Fitness Scale (IFIS), provides a valid approximation of objectively measured fitness levels. The IFIS has shown acceptable agreement with objective fitness measures across diverse populations, including children, adolescents, and young adults.^7,8^ Moreover, self-reported fitness assessed using the IFIS has shown associations with CVD risk factors that are comparable in both magnitude and direction to those observed with objective fitness measures.^7–9^ Despite these encouraging findings, most studies employing the IFIS have been conducted in populations with relatively low CVD risk, such as younger individuals, leaving a gap in evidence for high-risk populations, particularly those with T2DM. Given the elevated CVD risk inherent in T2DM,^10^ it is essential to investigate whether the IFIS can reliably evaluate fitness and its relationship with CVD risk factors in this population. Furthermore, no previous study has directly compared the predictive ability of the IFIS versus objectively measured fitness for CVD risk, including cardiometabolic and body composition outcomes, in any population.

Thus, the aims of this study were: 1) to investigate the validity of the IFIS self-reported physical fitness scores (i.e., overall and cardiorespiratory fitness) in assessing cardiorespiratory fitness considering the objectively measured 6-minute walk test as reference (criterion validity), and 2) to examine and compare the associations of the IFIS self-reported physical fitness scores and the 6-minute walk test with a wide range of cardiometabolic and body composition outcomes in adults with T2DM (construct validity).

## MATERIALS AND METHODS

### Study design and sample

This study utilizes cross-sectional data from the Evaluating the Prevalence and Severity Of MASLD (metabolic dysfunction-associated steatotic liver disease) In Primary care (EPSOMIP) study.^11^ The EPSOMIP study aimed to comprehensively explore the association between T2DM and metabolism-related conditions (i.e., MASLD, epicardial fat and CVD) using non-invasive advance techniques. It was conducted at four primary healthcare centers in the Swedish cities of Norrköping and Linköping, Sweden.

In the EPSOMIP protocol, inclusion criteria included a diagnosis of T2DM based on current guidelines and an age range of 35–75 years. Exclusion criteria encompassed previously diagnosed liver cirrhosis or other primary liver diseases aside from MASLD, contraindication for performing Magnetic Resonance Imaging (MRI), alcohol dependence, and inability to obtain Proton Density Fat Fraction (PDFF) measurements. Of the recruited participants, 18 withdrew consent, leaving 308 individuals enrolled in the EPSOMIP study. For the present analysis, after applying additional exclusion criteria, we included 282 participants with complete data on exposures, covariates, and at least one cardiometabolic or body composition outcome of interest. A flowchart of the study design is presented in **Figure 1**.

**Figure 1.**
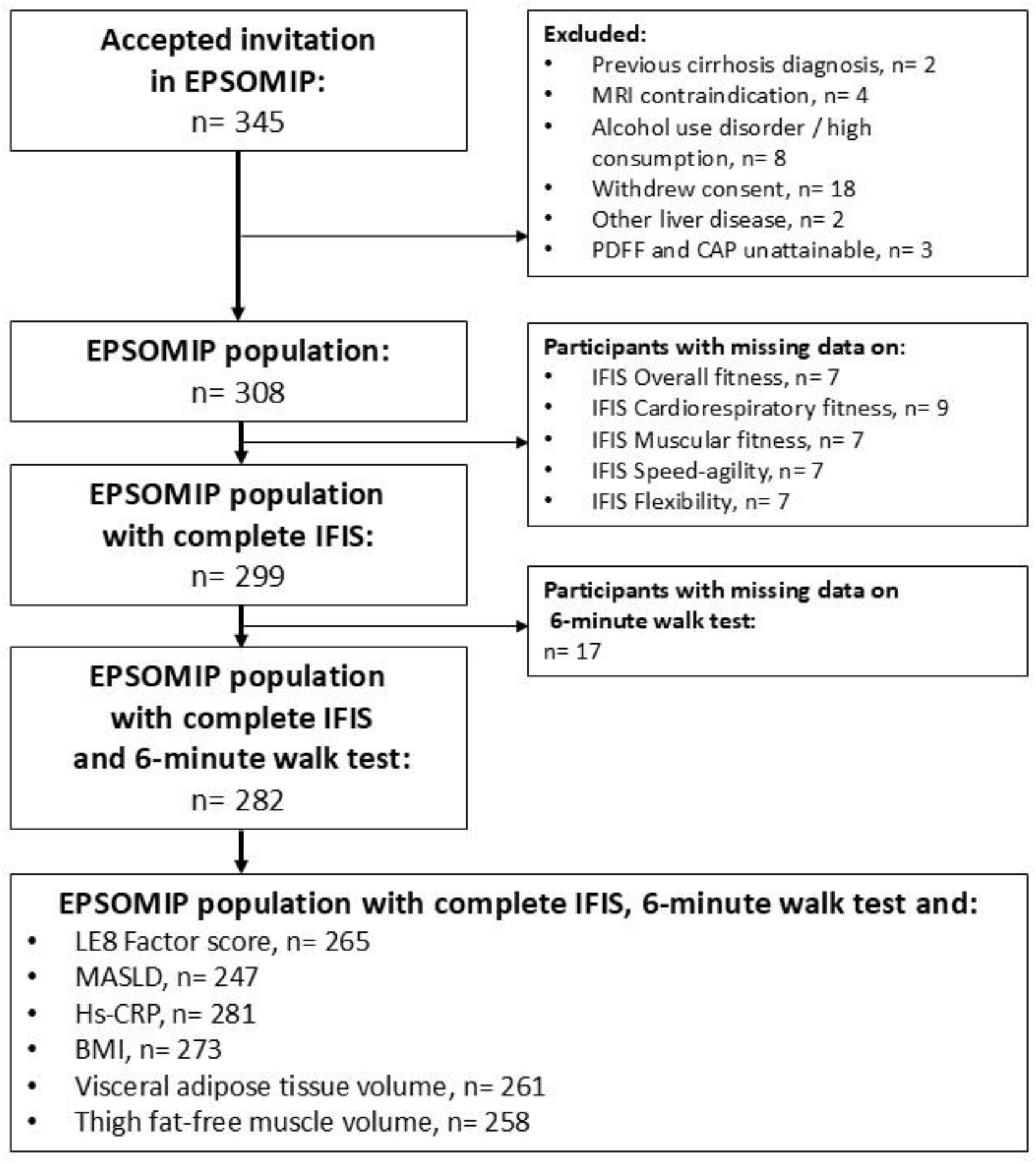
Flow chart of the study. BMI: body mass index, CAP: Controlled Attenuation Parameter, EPSOMIP: Evaluating the Prevalence and Severity Of MASLD in Primary Care, hs-CRP: high-sensitive C-reactive protein, IFIS: International Fitness Scale, LE8: Life’s Essential 8, MASLD: metabolic dysfunction-associated steatotic liver disease, MRI: Magnetic Resonance Imaging, PDFF: Proton Density Fat Fraction.

The Regional Ethical Board of Östergötland, Sweden, granted ethical approval for the EPSOMIP study (2018/176-31 and 2018/494-32), which also was prospectively registered (ClinicalTrials.gov identifier: NCT03864510). Written informed consent was obtained from each participant prior to inclusion, in accordance with the Declaration of Helsinki and Swedish accepted practices. Data collection and handling adhered to the General Data Protection Regulation (EU) 2016/679 and the standards outlined in Good Clinical Practice (ICH-GCP).

### Physical fitness

#### Self-reported physical fitness (IFIS)

Self-reported fitness was evaluated using the IFIS questionnaire.^8^ The questionnaire prompts participants to rate their fitness in five domains (overall fitness, cardiorespiratory fitness, muscular fitness, speed-agility, and flexibility) compared to peers of the same age. Each domain is rated on a Likert scale categorized as: “very poor” (1), “poor” (2), “average” (3), “good” (4), and “very good” (5). For this study, the Swedish version of the IFIS questionnaire was used (available at: https://profith.ugr.es/ifis) with minor modifications to adapt the language for an adult population.

#### Objectively measured cardiorespiratory fitness (6-minute walk test)

Cardiorespiratory fitness was objectively evaluated using the 6-minute walk test, a submaximal exercise test that measures the distance walked in meters over six minutes^8,12^ with proven validity across different populations.^13,14^ The test was conducted in a corridor with a flat, hard surface at the participants’ study centers as part of the EPSOMIP study. Participants were instructed to walk at a self-selected pace, allowing them to adjust their intensity and take breaks if needed. Standardized phrases of encouragement were provided at regular intervals to maintain motivation. The 6-minute walk test was used as a continuous variable and was further categorized as: <25^th^ percentile (197-488 meters), 25^th^-75^th^ percentile (489-595 meters) and >75^th^ percentile (596-794 meters).

### Cardiometabolic outcomes

The cardiovascular health score (Life’s Essential 8), liver fat, and high-sensitive C-reactive protein were considered relevant markers for assessing cardiometabolic outcomes. The cardiovascular health score effectively predicts CVD and mortality,^15,16^ while liver fat is linked to CVD and T2DM risk,^17^ and high-sensitive C-reactive protein reflects inflammation, a key driver of CVD conditions.^18^

Cardiovascular health was assessed using the Life’s Essential 8 Factor score, as defined by the American Heart Association.^19^ This composite score is derived from four factors closely associated with cardiovascular health: body mass index (BMI), blood lipids, blood pressure, and diabetes. The Factor score accounts for actual values of non-HDL cholesterol, blood pressure, and blood glucose, along with the use of medication for these factors, as well as values of BMI. A detailed description of the scoring methodology has been provided elsewhere.^20^ Briefly, and in line with the American Heart Association recommendations, each component was scored from 0 (poorest cardiovascular health) to 100 (best cardiovascular health). The Life’s Essential 8 Factor score was then calculated as the unweighted average of the four components, yielding an overall score ranging from 0 to 100. Additionally, a normalized Life’s Essential 8 Factor score was calculated by summing the Z-scores of the four components, which was then standardized into a single Z-score.

Liver fat (%) was quantified based on the hepatic triglyceride concentration, measured using PDFF by Magnetic Resonance Spectroscopy (MRS).^11^ Data were acquired using an 1.5T Achieva dStream MR scanner (Philips Healthcare, Best, The Netherlands).

High-sensitive C-reactive protein was measured in blood serum samples collected after an overnight fast. The lower detection limit for high-sensitive C-reactive protein was 0.15 mg/L; values below this threshold (n=7) were assigned a value of 0.10 mg/L.

### Body composition outcomes

Body composition markers like BMI, visceral fat, and muscle volume are strongly linked to T2DM and cardiovascular risk. BMI indicates obesity, visceral fat is closely associated to metabolic dysfunction,^21^ and low muscle mass is tied to impaired glucose metabolism and poorer health outcomes.^22,23^

BMI was measured using standardized procedures, calculated as weight divided by height squared, and expressed in units of kg/m^2^. Visceral adipose tissue volume (measured in liters, and henceforth referred as visceral adipose volume) and thigh fat-free muscle volume (measured in liters, and henceforth referred as muscle volume) were analyzed using AMRA® Researcher (AMRA Medical AB, Sweden) based on whole-body water-fat separated dual-echo Dixon MR images acquired using an 1.5T Achieva dStream MR scanner (Philips Healthcare, Best, The Netherlands).^24^ Visceral adipose volume was considered as a continuous variable, while for each participant in EPSOMIP, a sex, length, height, and BMI invariant muscle volume Z-score was also calculated.^25^ The muscle volume Z-score indicates how many standard deviations (SDs) the muscle volume of a participant deviates from what is expected given their sex and body size by comparing each participant to a matched group from the general population.

### Statistical analysis

In this study, we examined the distribution of continuous outcomes using both statistical (Kolmogorov–Smirnov test) and graphical (normal probability plots) methods. Variables that did not follow a normal distribution (i.e., high-sensitive C-reactive protein and liver fat) were natural-logarithmically transformed in the analysis. The validity of the IFIS scores in classifying T2DM participants into appropriate cardiorespiratory fitness levels was assessed using analysis of covariance (ANCOVA), both with and without adjustments for sex (categorical) and age (continuous). The 6-minute walk test served as the dependent variable, while the IFIS test categories were treated as fixed factors. Additionally, ANCOVA models adjusted for sex and age were used to investigate the associations between both self-reported and objectively measured fitness tests with cardiometabolic and body composition outcomes. For ANCOVA, all six outcomes were standardized to study-specific Z-scores to facilitate comparison between them (including standardization of the Life’s Essential 8 Factor Z-score and muscle volume Z-score).

The predictive ability of self-reported and objectively measured fitness tests for cardiometabolic and body composition outcomes was evaluated by comparing the areas under the curve (AUC) of unadjusted receiver operating characteristic (ROC) curves using DeLong tests. Additionally, predicted probabilities for health outcomes were generated using logistic regression models that incorporated self-reported or objective tests as predictors, adjusted for age and sex. The AUCs of those predicted probabilities were also compared using DeLong tests. For generating ROC curves, we selected clinically relevant outcomes as follows: poor cardiovascular health (<50 points in consonance with the recommendation of the American Heart Association),^19^ MASLD (≥5% of liver fat),^26^ high-risk high-sensitive C-reactive protein (>3.0 mg/L),^27^ obesity (BMI ≥30 kg/m^2^), high visceral adipose volume (≥75^th^ percentile), and low muscle volume <25^th^ percentile in a general population (< −0.68 SDs).^28^

To verify the robustness of our results, a series of sensitivity analyses were conducted: 1) excluding presumably low-intensity 6-minute walk tests defined as those with heart rate after the test below 60% of the predicted maximal heart rate (using Tanaka’s formula; predicted maximal heart rate=208−0.7×age), as those may indicate insufficient effort and affect test validity (n=79),^29^ 2) considering different cut-offs to dichotomize the outcomes in ROC curves, specifically all Z-scores for the outcomes were dichotomized at the median (0 indicating worse health, 1 indicating better health).

All statistical tests were two-sided and p<0.05 was considered statistically significant. Analyses were conducted using Stata V.18 (StataCorp 2021).

## RESULTS

### Participants’ characteristics

The characteristics of participants, stratified by the IFIS overall fitness, are presented in **Table 1**. Finally, 282 middle-aged participants with T2DM (37.9% women, mean age 63.6 ± 8.1 years) were included. Duration of T2DM was 8.4 ± 6.9 years, with 20.3% of participants receiving insulin and 87.5% receiving other treatments for T2DM.

**Table 1.** Clinical characteristics of the participants by IFIS overall fitness scores.

|  | All (n=282) |  | IFIS overall fitness (n=282) |  |  |
| --- | --- | --- | --- | --- | --- |
|  |  |  | Very poor/poor<br>n=37 | Average<br>n=132 | Good/very good<br>n=113 |
| **Sex (female, %)** | 282 | 107 (37.9) | 18 (48.6) | 51 (38.6) | 38 (33.6) |
| **Age (years)** | 282 | 63.6 ± 8.1 | 60.1 ± 10.0 | 63.4 ± 8.0 | 65.1 ± 7.1 |
| **Duration of T2DM (years)** | 279 | 8.4 ± 6.9 | 7.3 ± 4.6 | 8.4 ± 7.6 | 8.9 ± 6.8 |
| **Current use of diabetes medications** |  |  |  |  |  |
| Insulin | 271 | 55 (20.3) | 5 (14.3) | 32 (25.2) | 18 (16.5) |
| Metformin | 280 | 217 (77.5) | 27 (75.0) | 99 (75.6) | 91 (80.5) |
| Sulfonylurea | 274 | 25 (9.1) | 6 (16.7) | 12 (9.3) | 7 (6.4) |
| SGLT2-inhibitor | 272 | 80 (29.4) | 12 (32.4) | 42 (33.3) | 26 (23.9) |
| Incretin-based | 273 | 23 (8.4) | 4 (11.1) | 9 (7.0) | 10 (9.2) |
| **Smoking** | 282 |  |  |  |  |
| Never smoked |  | 138 (48.9) | 14 (37.8) | 59 (44.7) | 65 (57.5) |
| Current smoker |  | 11 (3.9) | 2 (5.4) | 4 (3.0) | 5 (4.4) |
| Ex-smoker |  | 133 (47.2) | 21 (56.8) | 69 (52.3) | 43 (38.1) |
| **6-minute walk test (meters)** | 282 | 538.9 ± 86.2 | 501.1 ± 79.0 | 533.1 ± 87.8 | 558.0 ± 81.9 |
| **6-minute walk test** | 282 |  |  |  |  |
| <25<sup>th</sup> percentile; <489 meters |  | 71 (25.2) | 14 (37.8) | 36 (27.3) | 21 (18.6) |
| 25-75<sup>th</sup> percentile; 489-595 meters |  | 142 (50.4) | 18 (48.6) | 66 (50.0) | 58 (51.3) |
| >75<sup>th</sup> percentile; ≥596 meters |  | 69 (24.5) | 5 (13.5) | 30 (22.7) | 34 (30.1) |
| **IFIS overall fitness** | 282 | 3.3 ± 0.8 | 1.9 ± 0.3 | 3.0 ± 0.0 | 4.1 ± 0.3 |
| **IFIS cardiorespiratory fitness** | 282 | 2.5 ± 0.9 | 1.4 ± 0.5 | 2.2 ± 0.7 | 3.2 ± 0.8 |
| **IFIS muscular fitness** | 282 | 3.3 ± 0.8 | 2.5 ± 0.9 | 3.2 ± 0.7 | 3.7 ± 0.6 |
| **IFIS speed-agility** | 282 | 2.9 ± 0.9 | 2.0 ± 0.8 | 2.7 ± 0.8 | 3.4 ± 0.7 |
| **IFIS flexibility** | 282 | 2.8 ± 0.9 | 1.9 ± 0.8 | 2.7 ± 0.7 | 3.3 ± 0.8 |
| **Cardiovascular health score** | 265 | 51.9 ± 12.8 | 43.8 ± 9.6 | 50.3 ± 13.3 | 56.3 ± 11.5 |
| **Liver fat (%)†** | 247 | 7.4 ± 11.9 | 15.7 ± 18.3 | 9.2 ± 11.7 | 5.2 ± 8.2 |
| **Hs-CRP (mg/L) †** | 281 | 1.2 ± 1.7 | 1.4 ± 2.4 | 1.3 ± 1.8 | 1.0 ± 1.7 |
| **BMI (kg/m2)** | 273 | 29.4 ± 4.5 | 32.7 ± 4.4 | 30.2 ± 4.5 | 27.5 ± 3.6 |
| **Visceral adipose tissue volume (L)** | 261 | 6.1 ± 2.6 | 7.6 ± 2.7 | 6.5 ± 2.5 | 5.2 ± 2.5 |
| **Thigh fat-free muscle volume (Z-score)** | 258 | 0.02 ± 1.05 | -0.04 ± 1.12 | -0.10 ± 1.01 | 0.16 ± 1.06 |

Categorical variables are depicted as frequency (percentages). Continuous variables normally and not normally distributed (marked with †) are depicted as mean ± standard deviation and median ± interquartile range, respectively.
Incretin-based medications refer to the use of glucagon-like peptide-1 receptor agonists (GLP-1 RAs) and/or inhibitors of the enzyme dipeptidyl peptidase-4 (DPP-4 inhibitors).
Tigh fat-free muscle volume (Z-score) is standardized by sex, length, height, and BMI from the general population.<sup>25</sup>
BMI: body mass index, hs-CRP: high-sensitivity C-reactive protein, IFIS: International Fitness Scale, SGLT2: sodium-glucose co-transporter 2 medication, T2DM: type 2 diabetes mellitus.

The mean in the 6-minute walk test was 538.9 ± 86.2 meters. For clinical outcomes, the mean cardiovascular health score, BMI, and visceral adipose volume were 51.9 points, 29.4 kg/m^2^, and 6.1 liters, respectively, while the median liver fat content and high-sensitive C-reactive protein levels were 7.4% and 1.2 mg/L, respectively.

**Supplementary Figure 1** illustrates the distribution of responses across all five IFIS scores. Overall, the distributions were relatively symmetric across the scores. However, in the IFIS cardiorespiratory test, participants more frequently scored “very poor” (14.9%) and less frequently scored “very good” (1.4%) compared to the other scores.

### Self-reported fitness versus objective cardiorespiratory fitness

Figure 2. illustrates the curvilinear dose-response associations between the IFIS overall fitness and cardiorespiratory fitness scores and the 6-minute walk test. Participants reporting better overall fitness generally achieved longer distances in the 6-minute walk test (p<0.001 for ANCOVA; see **Supplementary Table 1**). Specifically, participants reporting “very poor”, “poor”, “average”, “good” and “very good” on the IFIS overall fitness covered unadjusted mean distances of 508.2, 500.0, 533.1, 552.3 and 598.9 meters, respectively. A linear dose-response association was observed for IFIS cardiorespiratory fitness, with participants achieving unadjusted mean distances of 494.9, 531.5, 546.9, 579.6, and 624.5 meters, respectively, (p<0.001 for ANCOVA). A sensitivity analysis excluding those participants with a heart rate after 6-minute test below 60% of the predicted maximal heart rate (n=79) slightly attenuated the associations but did not alter the conclusions (**Supplementary Figure 2**).

**Figure 2.**
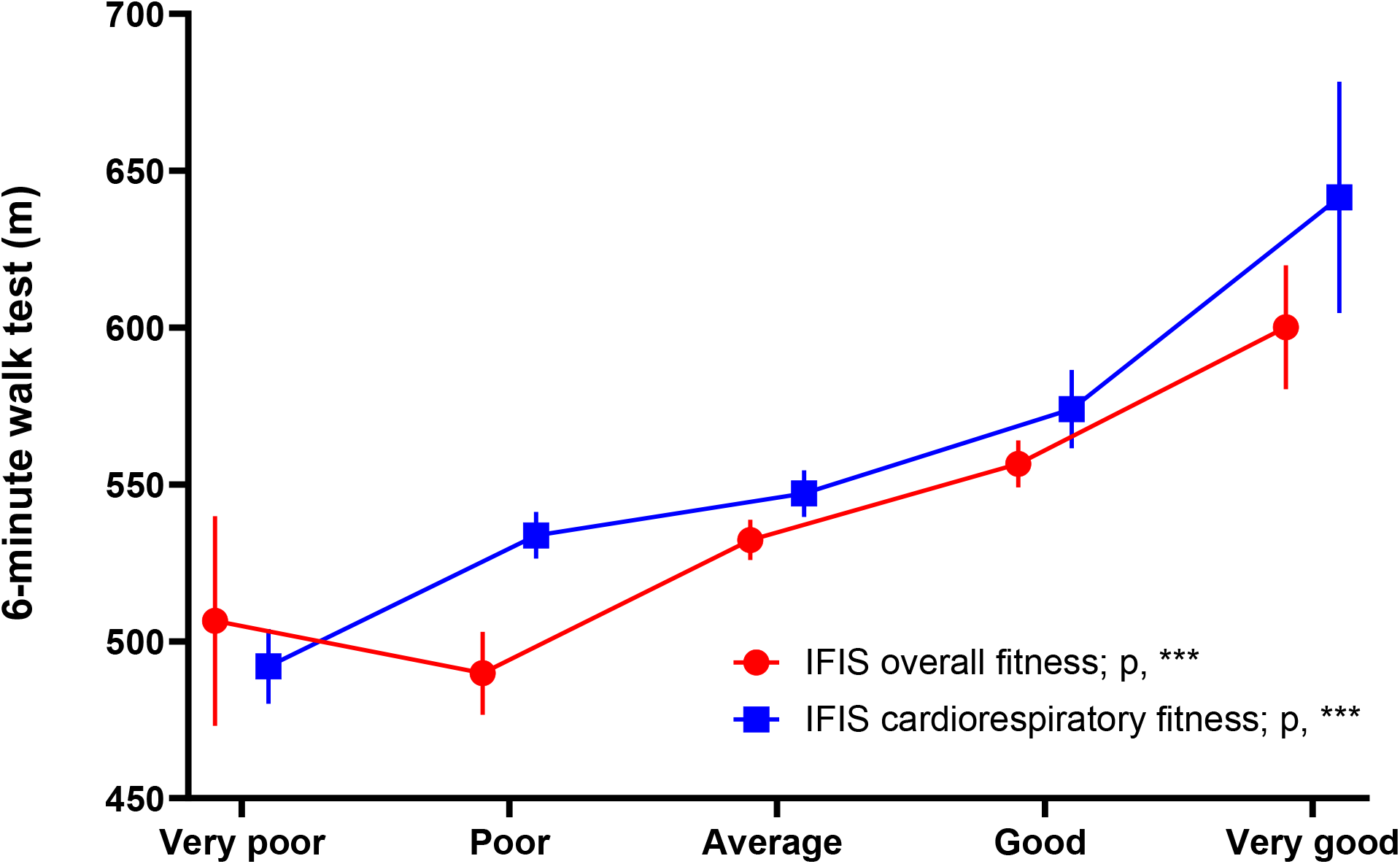
Comparison between self-reported (IFIS) overall and cardiorespiratory fitness versus objective (6-minute walk) cardiorespiratory fitness. The graphic depicts means with standard errors for the 6-minute walk test according to different levels of the IFIS scores. Analyses of covariance (ANCOVA) were adjusted for sex, and age. Significance level for each IFIS test is indicated as follows: ***, p<0.001; **, p<0.01; *, p<0.05; and ns, non-significant. IFIS: International Fitness Scale.

### Self-reported physical fitness in relation to cardiometabolic and body composition outcomes

**Figure 3** illustrates the associations between the IFIS scores (overall and cardiorespiratory fitness) and cardiometabolic (left panel) as well as body composition outcomes (right panel). Both IFIS scores showed significant associations with cardiometabolic and, particularly, body composition outcomes (p<0.05 for ANCOVA in all comparisons). In general, better IFIS scores were associated with higher cardiovascular health scores and muscle volume, alongside lower liver fat content, high-sensitive C-reactive protein, BMI, and visceral adipose volume. The patterns of association were largely similar to those observed with the 6-minute walk test (see **Supplementary Figure 3**).

**Figure 3.**
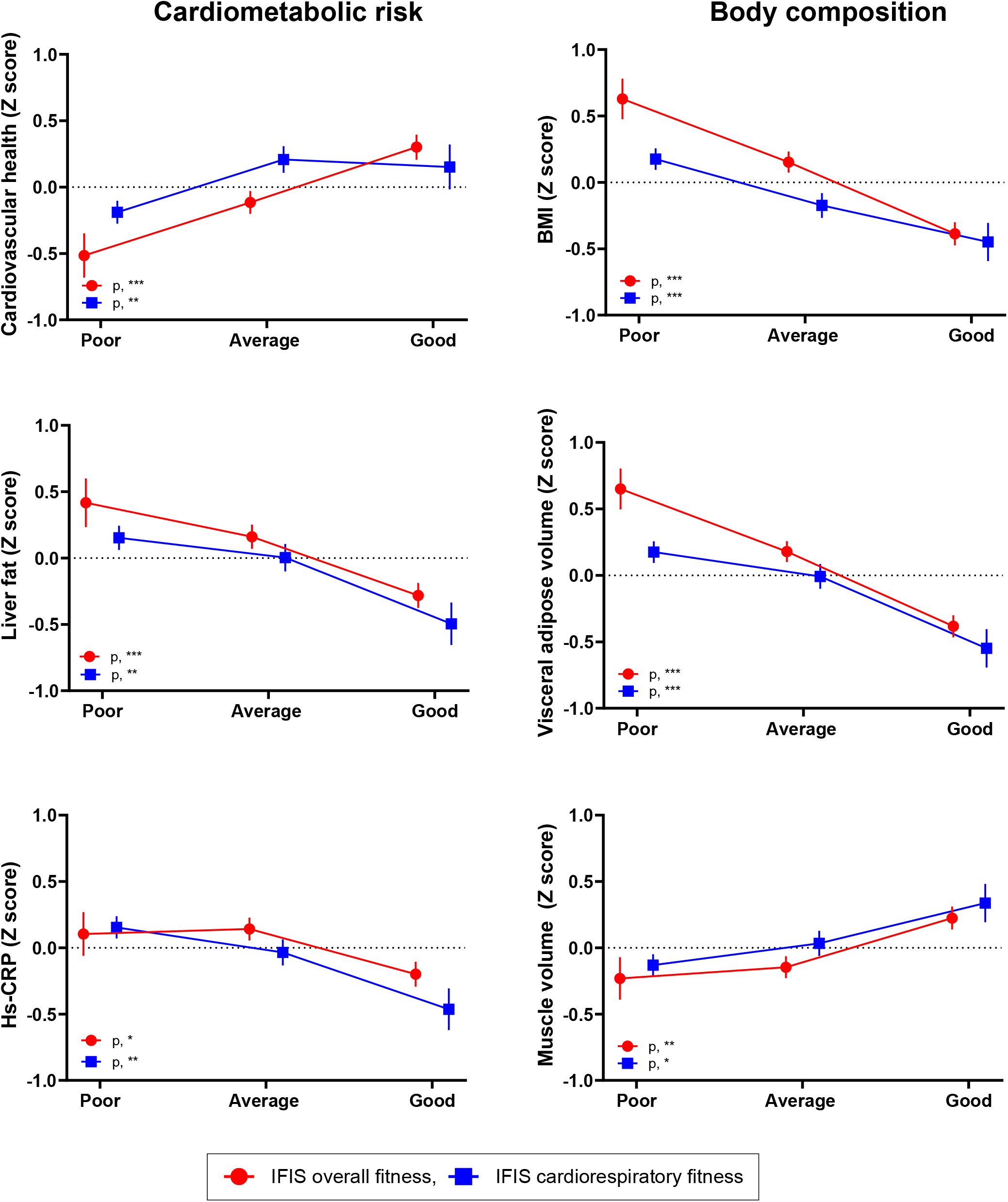
Differences in cardiometabolic risk and body composition outcomes according to categories of self-reported (IFIS) overall and cardiorespiratory fitness. The graphic depicts means and standard errors for the analyses of covariance (ANCOVA) adjusted for sex, and age. The variables liver fat and hs-CRP were natural-logarithmically transformed. All variables are presented as study-specific Z-scores (mean=0, standard deviation=1). IFIS scores are categorized as poor (very poor/poor), average and good (good/very good). Significance level for each IFIS test is indicated as follows: ***, p<0.001; **, p<0.01; *, p<0.05; and ns, non-significant. BMI: body mass index, hs-CRP: high-sensitive C-reactive protein, IFIS: International Fitness Scale.

However, the IFIS scores (overall and cardiorespiratory fitness) showed significant association for all cardiometabolic risk and body composition outcomes, while the 6-minute walk test did not show significant associations with liver fat and high-sensitive C-reactive protein (p>0.05 for ANCOVA).

**Supplementary Figure 4** depicts the associations between additional IFIS scores (muscular fitness, speed-agility and flexibility) and cardiometabolic and body composition outcomes. While IFIS speed-agility and IFIS flexibility showed significant associations with all cardiometabolic and body composition outcomes (p<0.05 for ANCOVA in all comparisons), IFIS muscular fitness only showed significant associations with visceral adipose volume and muscle volume.

### Predictive capacity of self-reported versus objective physical fitness for cardiometabolic and body composition outcomes

**Figure 4** depicts the predictive performance for the IFIS scores (overall and cardiorespiratory) compared to the 6-minute walk test for cardiometabolic and body composition outcomes. For cardiometabolic risk outcomes, the results showed that the predictive ability of the IFIS scores was at least as good as that of the 6-minute walk test. Notably, for predicting poor cardiovascular health, the IFIS overall fitness scores (AUC: 0.678, 95% CI: 0.618-0.737, p=0.031) outperformed the 6-minute walk test (AUC: 0.586, 95% CI: 0.517-0.654). Similarly, the IFIS overall fitness scores (AUC: 0.630, 95% CI: 0.563-0.696, p=0.016) outperformed the 6-minute walk test (AUC: 0.519, 95% CI: 0.443-0.595) for MASLD. For body composition outcomes, both IFIS scores demonstrated equivalent performance to the 6-minute walk test (p>0.05 for these AUC comparisons).

**Figure 4.**
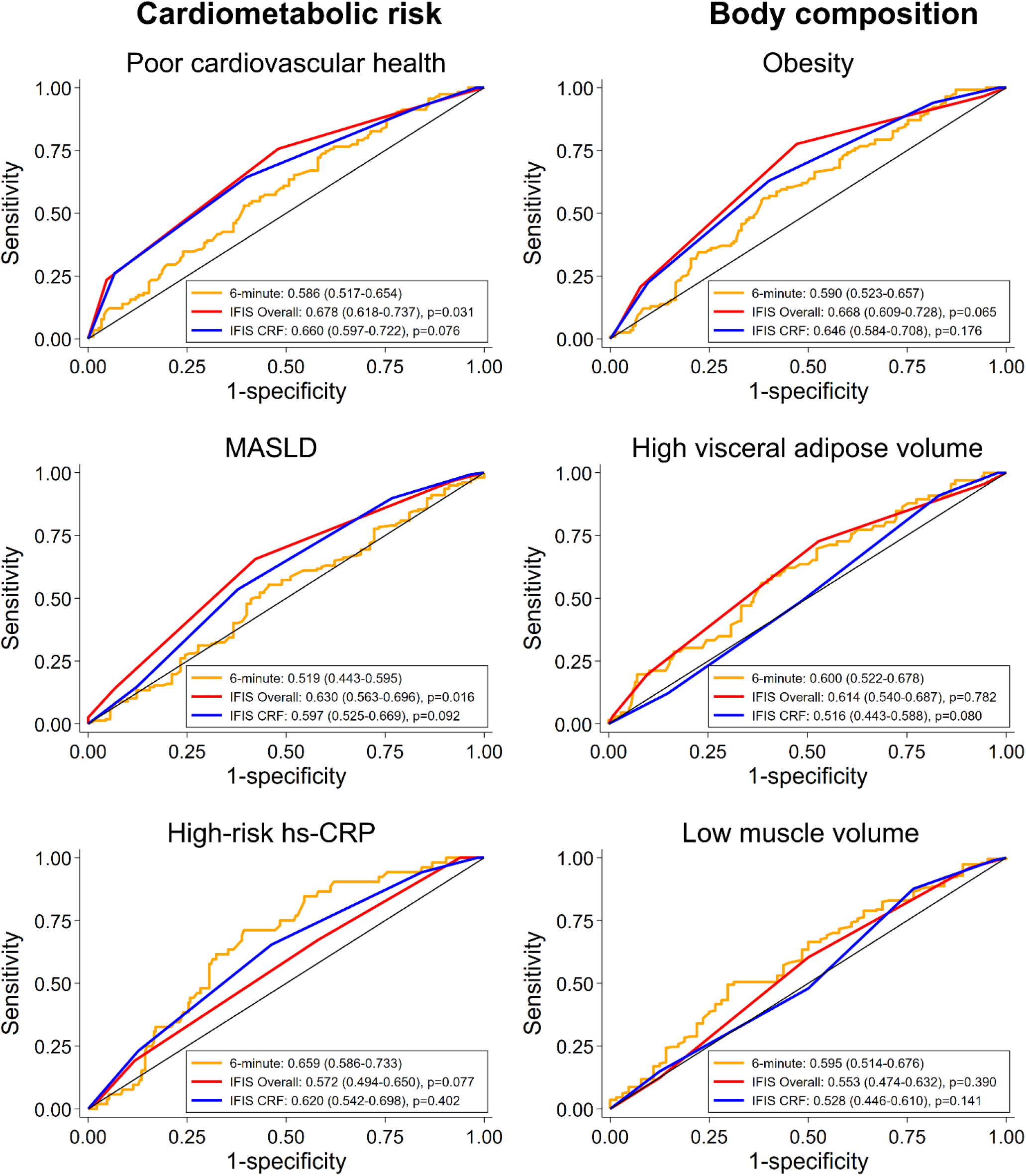
ROC curves comparing self-reported (IFIS) overall and cardiorespiratory fitness versus objective (6-minute walk) cardiorespiratory fitness for predicting cardiometabolic risk and body composition outcomes. The legends indicate the AUCs with corresponding 95% confidence intervals. The p-values represent comparisons of the AUCs between each specific IFIS test (IFIS overall and IFIS cardiorespiratory fitness) and the 6-minute walk test, performed using the DeLong test. AUC: area under curve, CRF: cardiorespiratory fitness, hs-CRP: high-sensitive C-reactive protein, IFIS: International Fitness Scale, MASLD: metabolic dysfunction-associated steatotic liver disease, ROC: receiver operating characteristic curve.

Overall, a sensitivity analysis using alternative cut-offs to dichotomize the outcomes (according to the median instead of standardized clinical outcomes) rendered similar conclusions, although the predictive capacity of the IFIS cardiorespiratory score to predict muscle volume was lower to that of the 6-minute walk test (instead of being similar, as observed in the main analysis) (**Supplementary Figure 5**).

**Supplementary Figure 6** depicts the predictive performance of the IFIS scores probabilities compared to the 6-minute walk test probabilities after adjusting for age and sex. It shows that the predictive ability of the IFIS scores was similar to that for the 6-minute walk test in all cardiovascular risk and body composition outcomes.

## DISCUSSION

This study supports the usefulness of self-reported fitness, measured with the IFIS, as a proxy of physical fitness in adults with T2DM. First, both the IFIS overall and the IFIS cardiorespiratory scores showed strong agreement with the objectively measured 6-minute walk test. Additionally, self-reported overall and cardiorespiratory fitness were both strongly linked to all cardiometabolic outcomes (i.e., cardiovascular health, liver fat, and high-sensitive C-reactive protein), and body composition outcomes (i.e., BMI, visceral adipose volume and muscle volume). Furthermore, ROC analysis revealed that the IFIS scores performed as well as, or better than the 6-minute walk test in predicting poor clinical cardiometabolic and body composition outcomes. Specifically, the IFIS overall score outperformed the 6-minute walk test for predicting poor cardiovascular health and MASLD.

To date, the IFIS has demonstrated good criterion validity for assessing cardiorespiratory fitness when compared with objectively measured cardiorespiratory fitness tests, such as the 6-minute walk test^8,29^ and the 20-meter shuttle run test.^30,31^ In line with this, our study found that the IFIS cardiorespiratory score obtained good agreement with the 6-minute walk test. To contextualize, our study found that the average distance between consecutive categories on the IFIS cardiorespiratory fitness was approximately 32 meters, a difference that is clinically meaningful as it exceeds the threshold of approximately 30 meters considered relevant for middle-aged patients with T2DM.^32^ Furthermore, the difference between the extremes (i.e., very poor versus very good cardiorespiratory fitness) was as high as 129.6 meters.

In addition to its criterion validity, several studies have established the construct validity of the IFIS, demonstrating associations with a range of health outcomes, including CVD risk factors (e.g., blood pressure, blood lipids, insulin resistance, inflammation, or metabolic syndrome) and adiposity indicators.^31^ However, existing evidence on the IFIS largely comes from studies on young and adolescent populations, with limited research exploring its validity in high CVD risk groups, particularly those with T2DM. By investigating the validity of the IFIS in middle-aged individuals with T2DM, who inherently face a heightened risk of CVD, this study may be of relevance for enhancing cardiovascular risk stratification.

In our study, IFIS cardiorespiratory fitness was strongly associated with both body composition and cardiometabolic outcomes in a population with T2DM. Notably, IFIS overall fitness, a simple question regarding one’s physical fitness relative to peers, also proved significant associations with all studied outcomes. Interestingly, our study also found that self-reported speed-agility and flexibility scores were strongly associated with cardiometabolic and body composition outcomes. In contrast, IFIS muscular fitness showed weaker associations, particularly with cardiometabolic outcomes and BMI. This finding aligns with previous literature across different populations,^7,9^ although such associations may vary depending on how muscular fitness is considered (i.e., absolute strength versus relative strength relative to body weight).^7^

To the best of our knowledge, no previous study has directly compared the predictive ability of IFIS and objectively measured fitness for cardiometabolic and body composition outcomes. Importantly, our ROC analyses showed that self-reported overall and cardiorespiratory fitness scores predicted cardiometabolic and body composition outcomes at least as well as the 6-minute walk test. In fact, we observed that the IFIS overall fitness score outperformed the 6-minute walk test to predict poor cardiovascular health and MALFD. Though the capacity of discrimination was moderate, the IFIS overall and cardiorespiratory fitness scores proved good construct validity to easily monitor not only physical fitness, but also cardiometabolic and body composition outcomes. Nevertheless, despite its potential, further studies are needed to evaluate the ability of the IFIS to predict hard clinical outcomes, including all-cause and CVD-related mortality and morbidity.

Cardiorespiratory fitness is supported by extensive epidemiological and clinical evidence as a strong predictor of mortality.^33^ Additionally, incorporating cardiorespiratory fitness into risk assessments alongside traditional factors significantly enhances the precision of risk reclassification for adverse outcomes or mortality.^33,34^ Recognizing the importance of this, the American Heart Association has identified cardiorespiratory fitness as a “vital clinical sign” and advocates for its routine assessment in clinical settings.^35^ In this sense, along with the proven criterion and construct validity, IFIS offers a highly feasible approach as it only requires a single self-reported response.

A strength of the study lies in the use of high-quality methods, such as body composition outcomes analyzed using fat and water separated MRI and MRS for liver fat, ensuring the comprehensive assessment of cardiometabolic and body composition outcomes. Furthermore, the EPSOMIP study utilized a population-based sample of patients with T2DM which increases the generalizability of the findings. However, several limitations should be noted. Test-retest reliability of IFIS was not assessed, though previous studies have demonstrated moderate to substantial reliability across populations,^7,8,36^ with minimal influence of learning or fatigue effects. Nonetheless, some heterogeneity in the reliability of the IFIS across populations must be acknowledged.^37^ Additionally, comparison with objective tests for muscular fitness, speed-agility, and flexibility was not included, which limits comparative analyses. However, cardiorespiratory fitness remains the physical fitness component most consistently associated with health outcomes.^35^ Finally, in our study, the 6-minute walk test serves as a practical proxy for laboratory-based cardiorespiratory tests. Despite its limitations, it has shown strong validity and is often the only feasible option in certain contexts.

Furthermore, sensitivity analyses excluding individuals with low-intensity 6-minute walk test potentially indicating insufficient effort yielded consistent results, further supporting the robustness of our findings.

## PERSPECTIVE

This study supports the utility of the IFIS test as a reliable tool for assessing physical fitness in middle-aged adults with T2DM, as it proved strong correlation with the 6-minute walk test. Furthermore, the IFIS predicted cardiometabolic risk and body composition outcomes at least as effectively as the 6-minute walk test. Thus, the IFIS provides an accessible, self-reported measure that may be easily integrated into clinical practice for physical fitness or cardiometabolic risk assessment in patients with an increased risk of CVD, such as individuals with T2DM. These findings build on previous research that has shown the potential of the IFIS to convey cardiovascular health in populations with low risk of CVD.^7,8,31,38–41^ However, further research is needed to assess its longitudinal validity and predictive power for morbidity and mortality outcomes. Additionally, ensuring demographic inclusivity in future studies is crucial to confirming its applicability across diverse populations. The inclusion of self-reported fitness assessments like the IFIS could improve preventive health strategies, particularly in Primary Care, where traditional fitness tests may be less feasible.

## CONCLUSIONS

Our findings demonstrate that self-reported fitness, as assessed by the IFIS, is a valid tool for evaluating physical fitness in individuals with T2DM, which may be important particularly in those settings where objective fitness tests may be impractical. In addition, IFIS overall and cardiorespiratory fitness scores generally convey cardiometabolic risk and body composition outcomes as least as good as the 6-minute walk test. In fact, IFIS overall fitness score was better than the 6-minute walk test for predicting cardiovascular health and MASLD. However, further studies are needed to examine the usefulness of the IFIS to predict major clinical outcomes such as all-cause and CVD-related mortality and morbidity.

## Data Availability

The dataset used and/or analyzed during the current study is available from the corresponding author upon reasonable request.

## Acknowledgments

We would like to thank Forum Östergötland for their support with study planning, legal guidance, and oversight of data quality. We also extend our gratitude to the coordinating study nurses, Carola Fagerström (Linköping) and Åsa Stahre Wiberg (Norrköping).

## Ethics approval and consent to participate

All recruitment processes and the collection of written informed consent were conducted in accordance with the World Medical Association Declaration of Helsinki (2018). Data collection adhered to the General Data Protection Regulation (EU) 2016/679 (GDPR) and complied with the International Conference on Harmonization—Good Clinical Practice (ICH-GCP) guidelines. The EPSONIP study received approval from the Regional Ethical Review Board (2018/176-31 and 2018/494-32) and is registered at clinicaltrials.gov (identifier: NCT03864510).

## Author’s contributions

Study concept and design: PH, AHA, KR, PN, ND, ODL, CJC, PL, SK, ME; Funding acquisition and supervision: PH, PN, ND, ODL, CJC, PL, SK, ME; Project administration and data curation: ME, FI, PL, SK; Patient recruitment: PN, FI, KR, SK, ME; Writing —original draft: AHA and PH; Statistical analysis: AHA, PH; Data analysis and interpretation: AHA, PH, KR; Constructive reviewing of the manuscript: KR, FI, CJC, NR, MF, ODL, PL, SK, ND, PN, ME. All authors read and approved of the final manuscript.

