## Supplementary material for "Self-Reported Fitness (IFIS) in Adults with Type 2 Diabetes Mellitus: Validity and Associations with Cardiometabolic and Body Composition Outcomes"

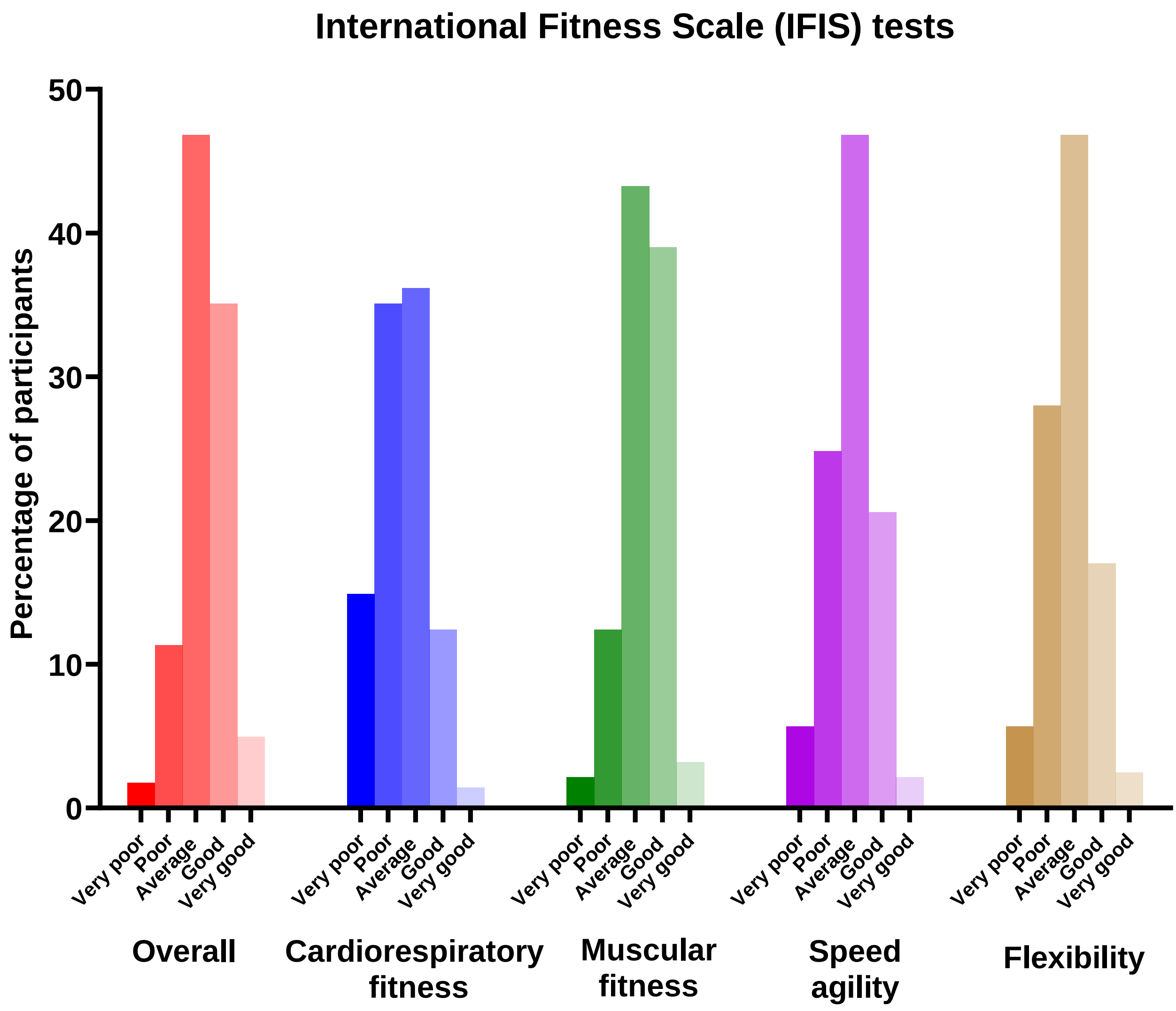


**Supplementary Figure 1. Distribution of responses to the five questions from the different self-reported (IFIS) scores.**
IFIS: International Fitness Scale.

### Main analysis vs excluding low-intensity 6-minute walk tests

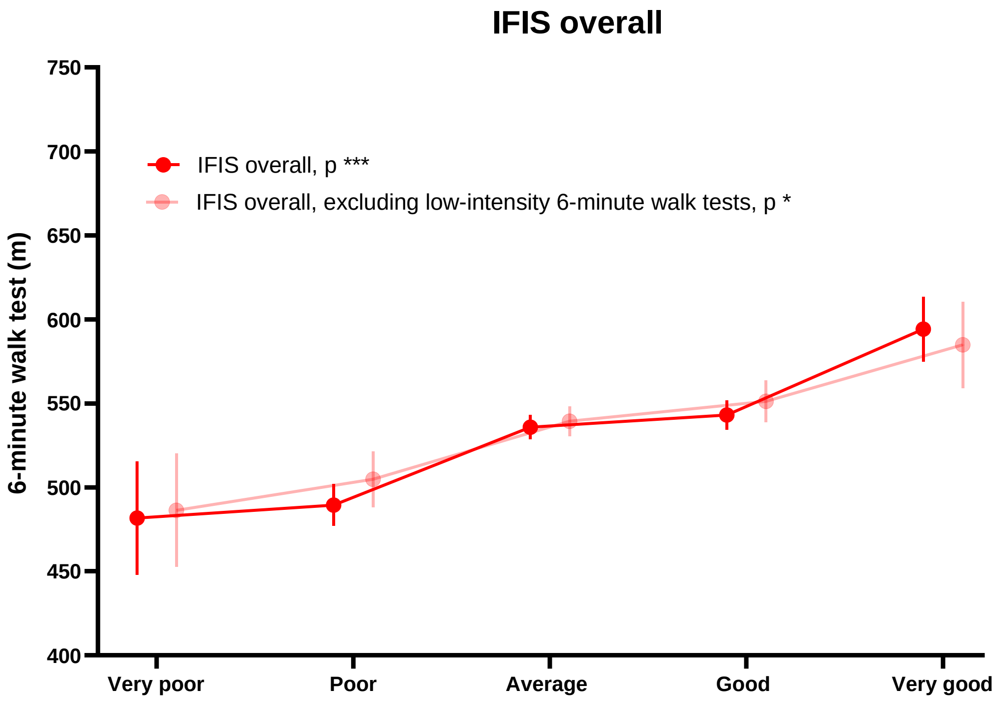


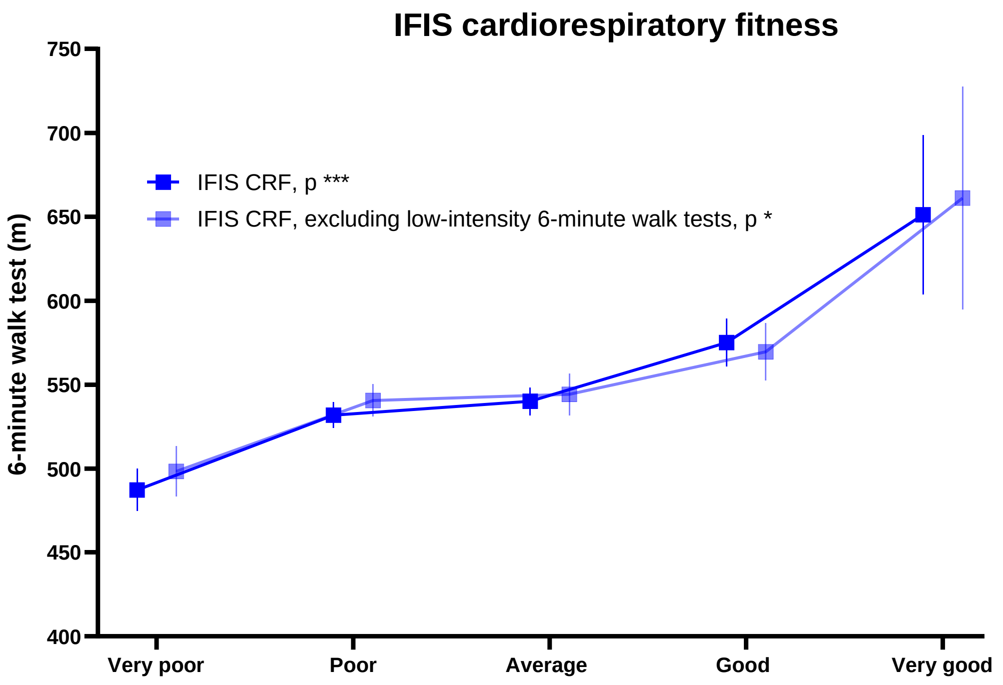


**Supplementary Figure 2. Sensitivity analysis comparing self-reported (IFIS) and objective (6-minute walk) physical fitness tests, including and excluding low-intensity 6-minute walk tests.**
The graphic depicts means with standard errors for the 6-minute walk test according to different levels of the IFIS scores. Analyses of covariance (ANCOVA) are adjusted for sex and age.
Presumably low-intensity 6-minute walk tests are considered those achieving less than 60% of the predicted maximal heart rate after test calculated with the Tanaka's formula. Significance level for each IFIS test is indicated as follows: ***, p<0.001; **, p<0.01; *, p<0.05; and ns, non-significant.
To allow comparability, population is restricted to those having information on the heart rate after the 6-minute walk test, IFIS overall and IFIS CRF (n=193), IFIS overall and IFIS CRF excluding low-intensity tests (n=114).
CRF: cardiorespiratory fitness, IFIS: International Fitness Scale.

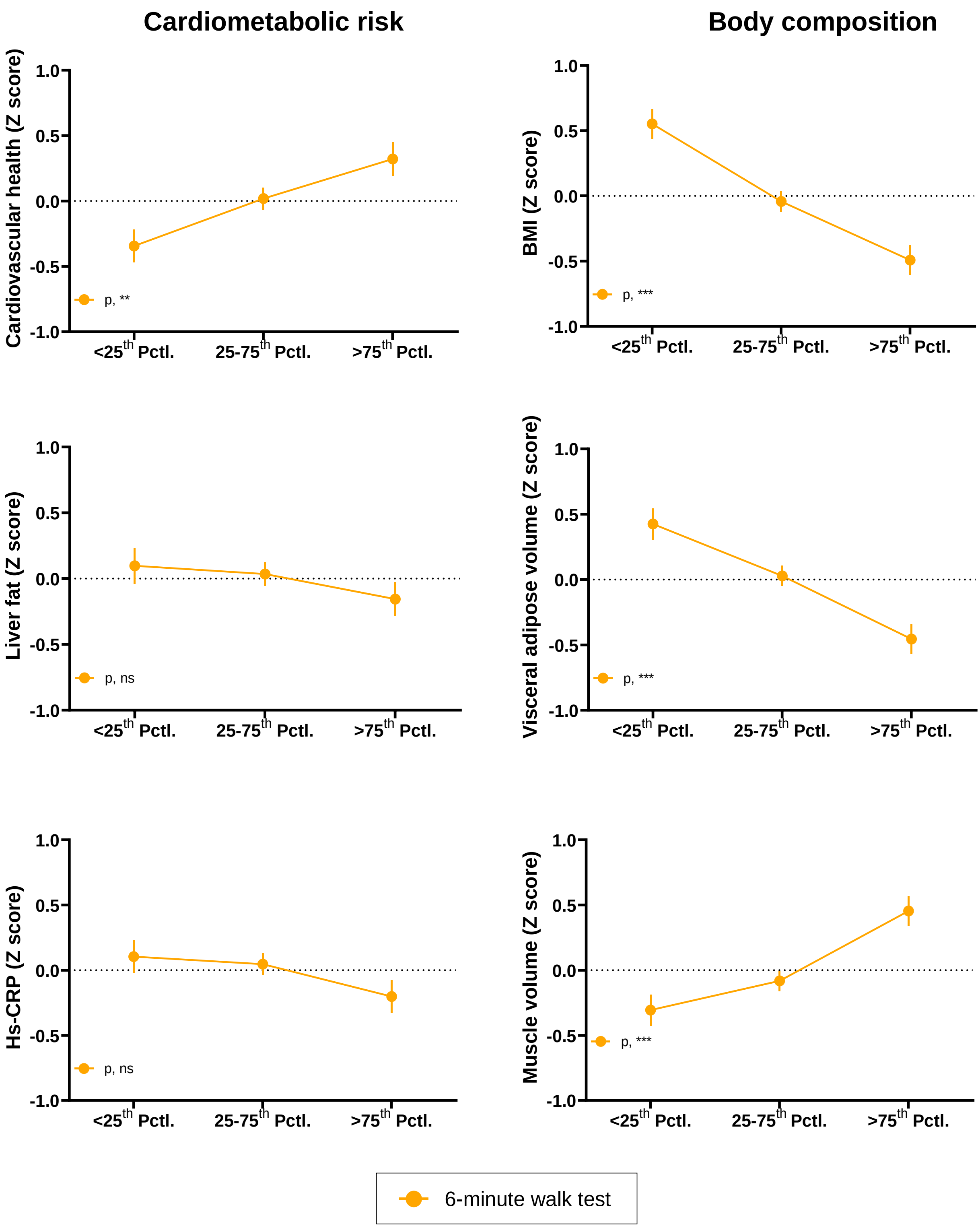


**Supplementary Figure 3. Differences in cardiometabolic risk and body composition outcomes according to percentiles groups of the 6-minute walk test.**

The graphic depicts means and standard errors for the analyses of covariance (ANCOVA) adjusted for sex and age. The variables liver fat and hs-CRP were natural-logarithmically transformed. All variables are presented as Z-scores (mean=0, standard deviation=1). The 6-minute walk tests are categorized as <25$^{th}$ percentile, 25-75$^{th}$ percentile, and >75$^{th}$ percentile. Significance level for each 6-minute walk test is indicated as follows: ***, p<0.001; **, p<0.01; *, p<0.05; and ns, non-significant.

BMI: body mass index, hs-CRP: high-sensitive C-reactive protein, Pctl: percentile.

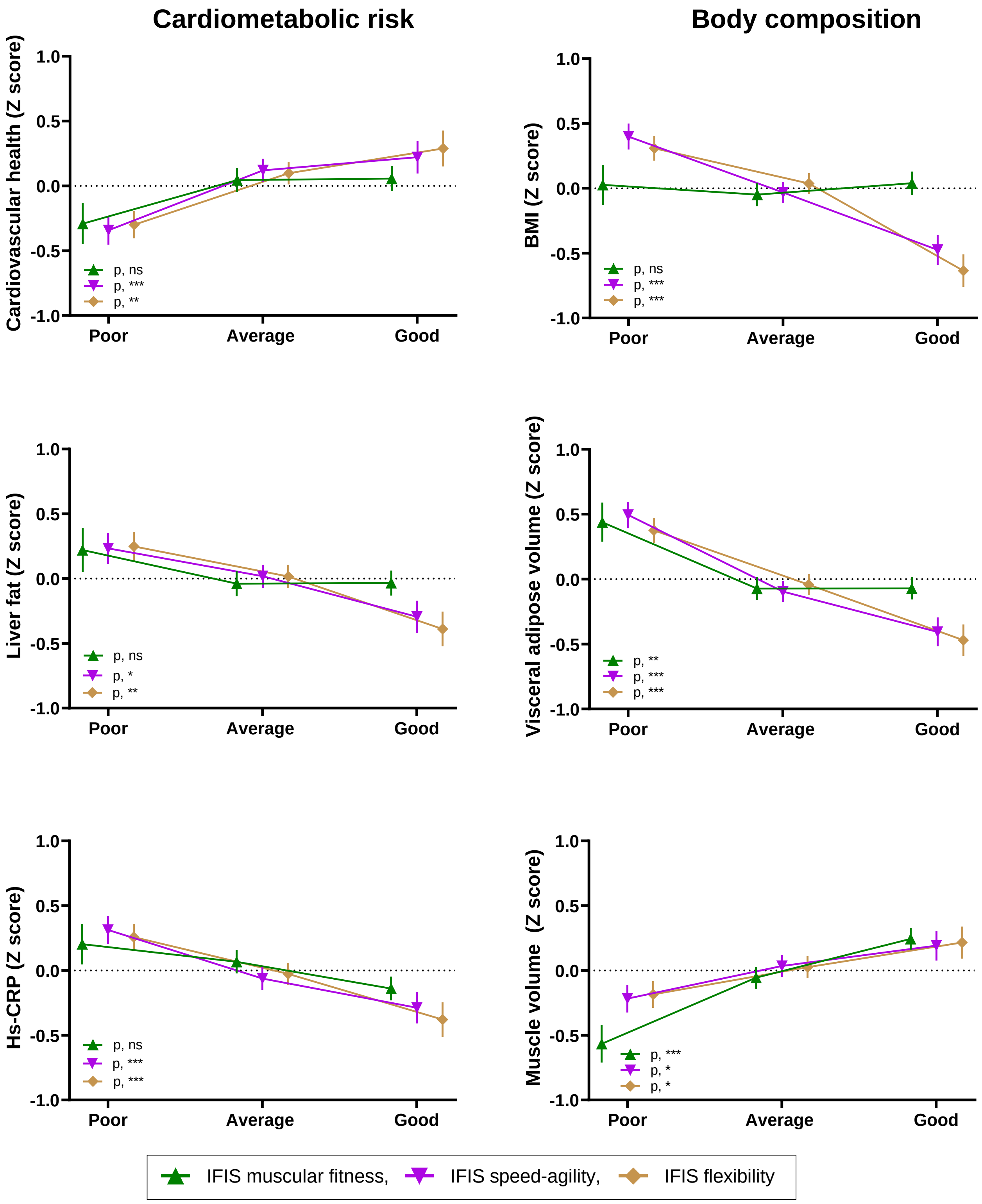


**Supplementary Figure 4. Differences in cardiometabolic risk and body composition outcomes according to categories of self-reported (IFIS) physical fitness scores.**

The graphic depicts means and standard errors for the analyses of covariance (ANCOVA) adjusted for sex and age. The variables liver fat and hs-CRP were natural-logarithmically transformed. All variables are presented as Z-scores (mean=0, standard deviation=1). IFIS scores are categorized as poor (very poor/poor), average and good (good/very good). Significance level for each IFIS test is indicated as follows: ***, p<0.001; **, p<0.01; *, p<0.05; and ns, non-significant.

BMI: body mass index, hs-CRP: high-sensitive C-reactive protein, IFIS: International Fitness Scale.

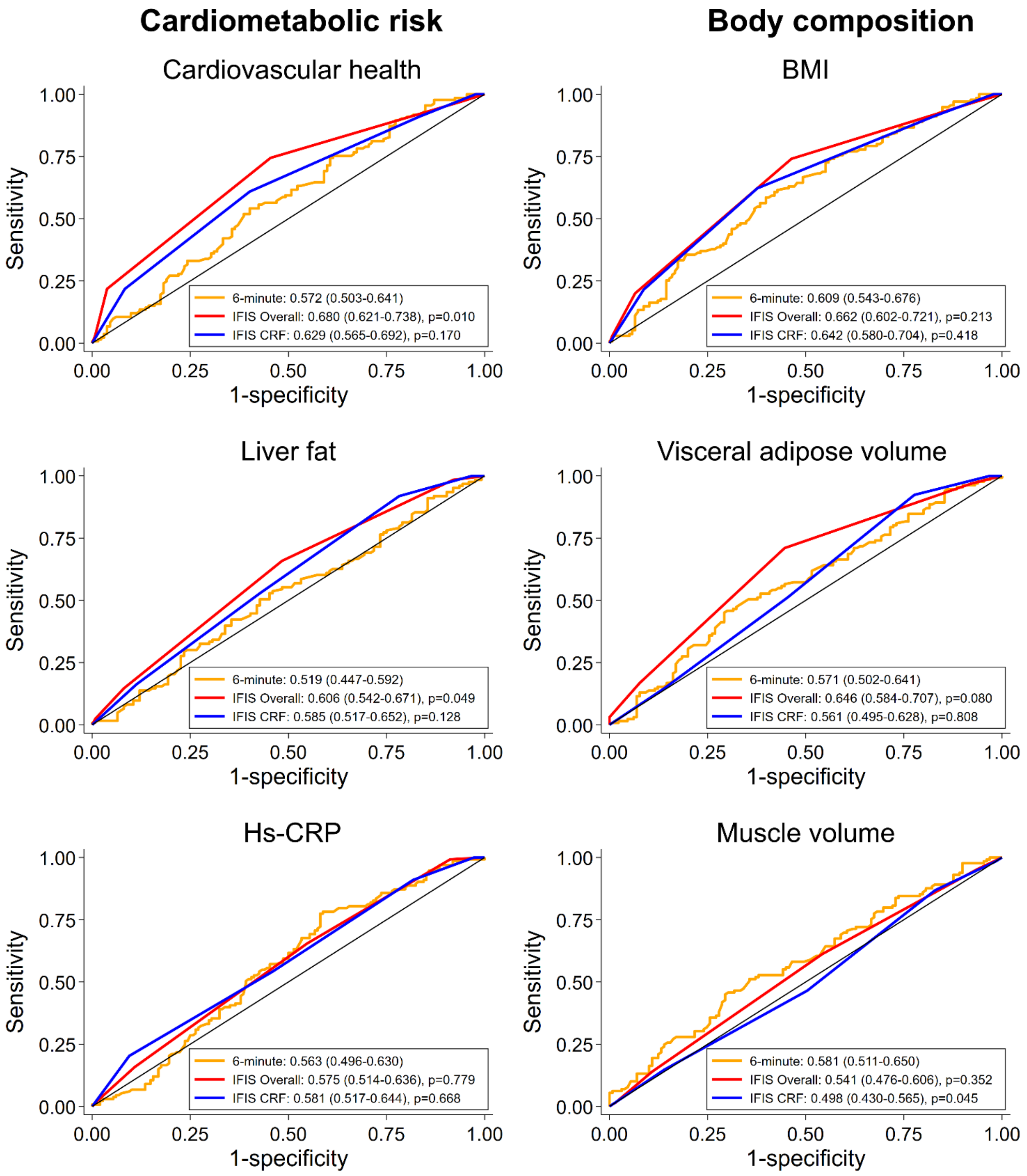


**Supplementary Figure 5. Sensitivity analysis, ROC curves comparing self-reported (IFIS) overall and cardiorespiratory fitness versus objective (6-minute walk) cardiorespiratory fitness for predicting cardiometabolic risk and body composition outcomes, considering a different categorization of the outcomes.**

The legends indicate the AUCs with corresponding 95% confidence intervals. The p-values represent comparisons of the AUCs between each specific IFIS test (IFIS overall and IFIS cardiorespiratory fitness) and the 6-minute walk test, performed using the DeLong test.

Outcomes are dichotomized at the median of the Z-scores (0 indicating worse health, 1 indicating better health).

AUC: area under curve, BMI: body mass index, CRF: cardiorespiratory fitness, hs-CRP: high-sensitive -reactive protein, IFIS: International Fitness Scale, ROC: receiver operating characteristic curve.

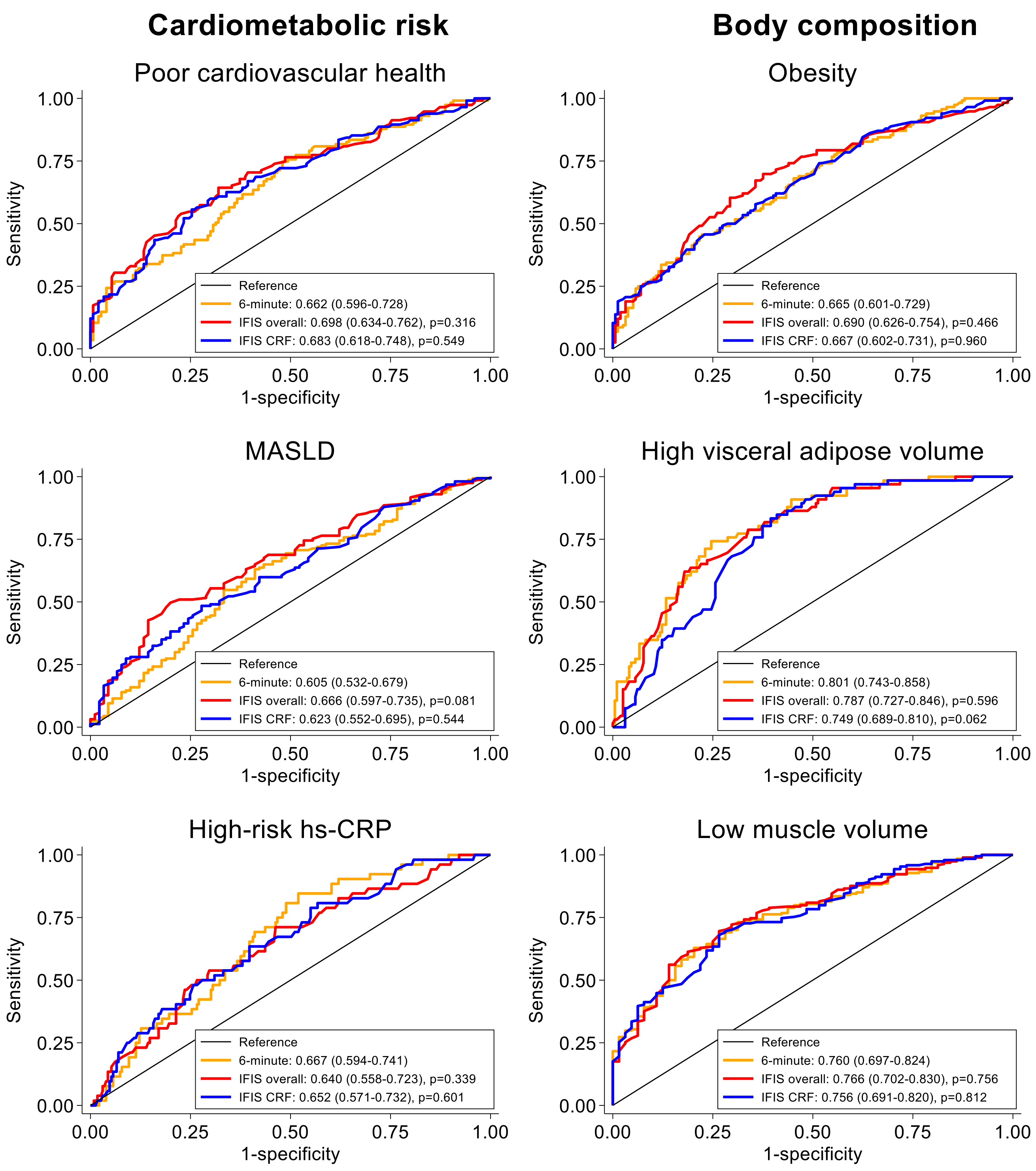


**Supplementary Figure 6. ROC curves for the probabilities of adjusted logistic models based on self-reported (IFIS) overall and cardiorespiratory fitness versus objective (6-minute walk) cardiorespiratory fitness for predicting cardiometabolic risk and body composition outcomes.**
The legends indicate AUCs with corresponding 95% confidence intervals. The p-values represent comparisons of the AUCs between the probabilities derived from the adjusted logistic models of the IFIS scores (IFIS overall and IFIS cardiorespiratory fitness) and the 6-minute walk test, performed using the DeLong test. All models are adjusted for age (continuous) and sex (female/male).
AUC: area under curve, CRF: cardiorespiratory fitness, hs-CRP: high-sensitive C-reactive protein, IFIS: International Fitness Scale, MASLD: metabolic dysfunction-associated steatotic liver disease, ROC: receiver operating characteristic curve.

**Supplementary Table 1. Differences in the 6-minute walk test according to categories of self-reported (IFIS) physical fitness scores.**

| | 6-minute walk test | | | | | |
|---|---|---|---|---|---|---|
| | Unadjusted | | | Adjusted | | |
| | Category | Mean | 95% CI | Category | Mean | 95% CI |
| **IFIS overall fitness** | Very poor [-] | 508.2 | (434.1, 582.3) | Very poor [-] | 506.5 | (440.8, 572.2) |
| | Poor [G, VG] | 500.0 | (470.7, 529.3) | Poor [A, G, VG] | 489.8 | (463.9, 515.8) |
| | Average [VG] | 533.1 | (518.7, 547.6) | Average [P, VG] | 532.3 | (519.7, 545.0) |
| | Good [P] | 552.3 | (535.6, 568.9) | Good [P] | 556.6 | (541.8, 571.3) |
| | Very good [P, A] | 598.9 | (554.6, 643.1) | Very good [P, A] | 600.1 | (561.2, 638.9) |
| **IFIS cardiorespiratory fitness** | Very poor [A, G, VG] | 494.9 | (469.7, 520.1) | Very poor [P, A, G, VG] | 492.0 | (468.7, 515.4) |
| | Poor [G] | 531.5 | (515.1, 547.9) | Poor [VP, G, VG] | 533.8 | (519.2, 548.4) |
| | Average [VP] | 546.9 | (530.7, 563.1) | Average [VP] | 547.1 | (532.5, 561.7) |
| | Good [VP, P] | 579.6 | (552.0, 607.2) | Good [VP] | 574.0 | (549.4, 598.7) |
| | Very good [VP] | 624.5 | (542.9, 706.1) | Very good [VP, P] | 641.5 | (568.9, 714.1) |

Data are presented as means and 95% confidence intervals. Adjusted models are adjusted by age and sex. Superscripts indicate statistically significant Tukey's pairwise comparisons ($p<0.05$) for the means of the 6-minute walk test across categories of the IFIS scores: VP (Very poor), P (Poor), A (Average), G (Good), and VG (Very good). For example, in IFIS overall fitness, for the unadjusted model, those rating their overall fitness as "Poor" had significant differences in the 6-minute walk test compared to those rating their overall fitness as "Good" or "Very good".
CI: confidence interval, IFIS: International Fitness Scale.

**Supplementary Table 2. Differences in cardiometabolic outcomes according to categories of self-reported (IFIS) physical fitness scores.**

| | Unadjusted | | | Adjusted | | |
|---|---|---|---|---|---|---|
| | **Category** | **Mean** | **95% CI** | **Category** | **Mean** | **95% CI** |
| **Cardiovascular health (Z-score)** | | | | | | |
| **IFIS overall fitness** | Poor $^{-,G}$ | -0.538 | (-0.863, -0.214) | Poor $^{-,G}$ | -0.515 | (-0.844, -0.186) |
| | Average $^{-,G}$ | -0.120 | (-0.289, 0.050) | Average $^{-,G}$ | -0.115 | (-0.284, 0.054) |
| | Good $^{P,A}$ | 0.314 | (0.130, 0.497) | Good $^{P,A}$ | 0.301 | (0.115, 0.487) |
| **IFIS cardiorespiratory fitness** | Poor $^{A,-}$ | -0.190 | (-0.357, -0.022) | Poor $^{A,-}$ | -0.189 | (-0.359, -0.020) |
| | Average $^{P,-}$ | 0.208 | (0.011, 0.405) | Average $^{P,-}$ | 0.208 | (0.010, 0.406) |
| | Good $^{-,-}$ | 0.154 | (-0.178, 0.487) | Good $^{-,-}$ | 0.152 | (-0.181, 0.485) |
| **IFIS muscular fitness** | Poor $^{-,-}$ | -0.289 | (-0.604, 0.025) | Poor $^{-,-}$ | -0.291 | (-0.606, 0.023) |
| | Average $^{-,-}$ | 0.048 | (-0.135, 0.231) | Average $^{-,-}$ | 0.045 | (-0.137, 0.227) |
| | Good $^{-,-}$ | 0.052 | (-0.135, 0.238) | Good $^{-,-}$ | 0.056 | (-0.129, 0.241) |
| **IFIS speed-agility** | Poor $^{A,G}$ | -0.374 | (-0.585, -0.162) | Poor $^{A,G}$ | -0.342 | (-0.557, -0.128) |
| | Average $^{P,-}$ | 0.137 | (-0.035, 0.309) | Average $^{P,-}$ | 0.120 | (-0.053, 0.294) |
| | Good $^{P,-}$ | 0.230 | (-0.017, 0.476) | Good $^{P,-}$ | 0.222 | (-0.025, 0.469) |
| **IFIS flexibility** | Poor $^{A,G}$ | -0.321 | (-0.523, -0.118) | Poor $^{A,G}$ | -0.299 | (-0.502, -0.096) |
| | Average $^{P,-}$ | 0.111 | (-0.062, 0.283) | Average $^{P,-}$ | 0.098 | (-0.074, 0.271) |
| | Good $^{P,-}$ | 0.297 | (0.028, 0.566) | Good $^{P,-}$ | 0.288 | (0.020, 0.557) |
| **Liver fat (Z-score)** | | | | | | |
| **IFIS overall fitness** | Poor $^{-,G}$ | 0.458 | (0.099, 0.817) | Poor $^{-,G}$ | 0.417 | (0.056, 0.777) |
| | Average $^{-,G}$ | 0.170 | (-0.009, 0.349) | Average $^{-,G}$ | 0.161 | (-0.018, 0.339) |
| | Good $^{P,A}$ | -0.302 | (-0.487, -0.118) | Good $^{P,A}$ | -0.281 | (-0.467, -0.096) |
| **IFIS cardiorespiratory fitness** | Poor $^{-,G}$ | 0.177 | (0.000, 0.354) | Poor $^{-,G}$ | 0.153 | (-0.027, 0.333) |
| | Average $^{-,G}$ | -0.027 | (-0.228, 0.173) | Average $^{-,G}$ | 0.003 | (-0.200, 0.207) |
| | Good $^{P,A}$ | -0.497 | (-0.814, -0.181) | Good $^{P,A}$ | -0.495 | (-0.810, -0.180) |
| **IFIS muscular fitness** | Poor $^{-,-}$ | 0.249 | (-0.084, 0.582) | Poor $^{-,-}$ | 0.221 | (-0.111, 0.554) |
| | Average $^{-,-}$ | -0.058 | (-0.250, 0.133) | Average $^{-,-}$ | -0.039 | (-0.229, 0.151) |
| | Good $^{-,-}$ | -0.024 | (-0.215, 0.167) | Good $^{-,-}$ | -0.034 | (-0.224, 0.156) |
| **IFIS speed-agility** | Poor $^{-,G}$ | 0.250 | (0.015, 0.485) | Poor $^{-,G}$ | 0.233 | (-0.002, 0.468) |
| | Average $^{-,-}$ | 0.006 | (-0.172, 0.185) | Average $^{-,-}$ | 0.018 | (-0.160, 0.196) |
| | Good $^{P,-}$ | -0.291 | (-0.539, -0.042) | Good $^{P,-}$ | -0.295 | (-0.541, -0.048) |
| **IFIS flexibility** | Poor $^{-,G}$ | 0.241 | (0.021, 0.461) | Poor $^{-,G}$ | 0.248 | (0.030, 0.466) |
| | Average $^{-,G}$ | 0.013 | (-0.166, 0.192) | Average $^{-,G}$ | 0.016 | (-0.161, 0.193) |
| | Good $^{P,A}$ | -0.372 | (-0.635, -0.110) | Good $^{P,A}$ | -0.389 | (-0.649, -0.129) |
| **Hs-CRP (Z-score)** | | | | | | |
| **IFIS overall fitness** | Poor $^{-,-}$ | 0.133 | (-0.187, 0.453) | Poor $^{-,-}$ | 0.104 | (-0.220, 0.428) |
| | Average $^{-,G}$ | 0.143 | (-0.027, 0.313) | Average $^{-,G}$ | 0.142 | (-0.028, 0.312) |
| | Good $^{-,A}$ | -0.209 | (-0.392, -0.026) | Good $^{-,A}$ | -0.199 | (-0.383, -0.014) |
| **IFIS cardiorespiratory fitness** | Poor $^{-,G}$ | 0.176 | (0.013, 0.339) | Poor $^{-,G}$ | 0.154 | (-0.011, 0.320) |
| | Average $^{-,-}$ | -0.058 | (-0.249, 0.133) | Average $^{-,-}$ | -0.035 | (-0.228, 0.159) |
| | Good $^{P,-}$ | -0.481 | (-0.789, -0.172) | Good $^{P,-}$ | -0.463 | (-0.772, -0.154) |
| **IFIS muscular fitness** | Poor $^{-,-}$ | 0.237 | (-0.069, 0.542) | Poor $^{-,-}$ | 0.203 | (-0.103, 0.510) |
| | Average $^{-,-}$ | 0.057 | (-0.120, 0.234) | Average $^{-,-}$ | 0.067 | (-0.110, 0.244) |
| | Good $^{-,-}$ | -0.141 | (-0.321, 0.039) | Good $^{-,-}$ | -0.140 | (-0.319, 0.040) |
| **IFIS speed-agility** | Poor $^{A,G}$ | 0.299 | (0.090, 0.508) | Poor $^{A,G}$ | 0.313 | (0.102, 0.524) |
| | Average $^{P,-}$ | -0.057 | (-0.225, 0.111) | Average $^{P,-}$ | -0.062 | (-0.230, 0.106) |
| | Good $^{P,-}$ | -0.279 | (-0.520, -0.038) | Good $^{P,-}$ | -0.288 | (-0.527, -0.048) |
| **IFIS flexibility** | Poor $^{-,G}$ | 0.241 | (0.043, 0.439) | Poor $^{-,G}$ | 0.257 | (0.059, 0.455) |
| | Average $^{-,-}$ | -0.025 | (-0.194, 0.144) | Average $^{-,-}$ | -0.027 | (-0.195, 0.141) |
| | Good $^{P,-}$ | -0.357 | (-0.617, -0.097) | Good $^{P,-}$ | -0.379 | (-0.638, -0.120) |

Data are presented as mean and 95% confidence intervals. Adjusted models are adjusted by age and sex. Superscripts indicate statistically significant Tukey's pairwise comparisons ($p<0.05$) for the Z-scores of cardiometabolic outcomes across categories of the IFIS scores: P (Poor), A (Average), and G (Good). For example, in IFIS overall fitness, for the unadjusted model, those rating their overall fitness as "Poor" had significant differences in the Z-score for cardiovascular health compared to those rating their overall fitness as "Good".

CI: confidence interval, hs-CRP: high-sensitivity C-reactive protein, IFIS: International Fitness Scale.

**Supplementary Table 3. Differences in body composition outcomes according to categories of self-reported (IFIS) physical fitness scores.**

| | Unadjusted | | | Adjusted | | |
|---|---|---|---|---|---|---|
| | Category | Mean | 95% CI | Category | Mean | 95% CI |
| **Body mass index (Z-score)** | | | | | | |
| **IFIS overall fitness** | Poor [A,G] | 0.731 | (0.429, 1.033) | Poor [A,G] | 0.628 | (0.328, 0.928) |
| | Average [P,G] | 0.165 | (0.005, 0.325) | Average [P,G] | 0.153 | (-0.004, 0.309) |
| | Good [P,A] | -0.435 | (-0.609, -0.261) | Good [P,A] | -0.387 | (-0.558, -0.216) |
| **IFIS cardiorespiratory fitness** | Poor [A,G] | 0.286 | (0.124, 0.447) | Poor [A,G] | 0.248 | (0.088, 0.407) |
| | Average [P,-] | -0.215 | (-0.403, -0.028) | Average [P,-] | -0.174 | (-0.358, 0.011) |
| | Good [P,-] | -0.475 | (-0.789, -0.160) | Good [P,-] | -0.448 | (-0.754, -0.143) |
| **IFIS muscular fitness** | Poor [-,-] | 0.065 | (-0.243, 0.373) | Poor [-,-] | 0.027 | (-0.271, 0.324) |
| | Average [-,-] | -0.072 | (-0.255, 0.111) | Average [-,-] | -0.049 | (-0.225, 0.127) |
| | Good [-,-] | 0.049 | (-0.134, 0.232) | Good [-,-] | 0.039 | (-0.136, 0.215) |
| **IFIS speed-agility** | Poor [A,G] | 0.461 | (0.260, 0.661) | Poor [A,G] | 0.399 | (0.203, 0.596) |
| | Average [P,G] | -0.072 | (-0.238, 0.094) | Average [P,G] | -0.032 | (-0.193, 0.129) |
| | Good [P,A] | -0.478 | (-0.712, -0.245) | Good [P,A] | -0.475 | (-0.701, -0.250) |
| **IFIS flexibility** | Poor [A,G] | 0.337 | (0.146, 0.529) | Poor [-,G] | 0.309 | (0.124, 0.493) |
| | Average [P,G] | 0.011 | (-0.154, 0.177) | Average [-,G] | 0.037 | (-0.122, 0.196) |
| | Good [P,A] | -0.625 | (-0.880, -0.369) | Good [P,A] | -0.635 | (-0.880, -0.391) |
| **Visceral adipose volume (Z-score)** | | | | | | |
| **IFIS overall fitness** | Poor [-,G] | 0.560 | (0.228, 0.892) | Poor [A,G] | 0.650 | (0.348, 0.953) |
| | Average [-,G] | 0.159 | (-0.013, 0.331) | Average [P,G] | 0.179 | (0.024, 0.334) |
| | Good [P,A] | -0.335 | (-0.514, -0.156) | Good [P,A] | -0.383 | (-0.545, -0.220) |
| **IFIS cardiorespiratory fitness** | Poor [-,G] | 0.064 | (-0.108, 0.237) | Poor [-,G] | 0.176 | (0.015, 0.336) |
| | Average [-,G] | 0.108 | (-0.089, 0.306) | Average [-,G] | -0.008 | (-0.191, 0.176) |
| | Good [P,A] | -0.473 | (-0.784, -0.163) | Good [P,A] | -0.549 | (-0.832, -0.265) |
| **IFIS muscular fitness** | Poor [-,-] | 0.339 | (0.017, 0.661) | Poor [A,G] | 0.438 | (0.142, 0.735) |
| | Average [-,-] | -0.051 | (-0.237, 0.135) | Average [P,-] | -0.073 | (-0.244, 0.097) |
| | Good [-,-] | -0.061 | (-0.245, 0.123) | Good [P,-] | -0.071 | (-0.241, 0.098) |
| **IFIS speed-agility** | Poor [A,G] | 0.526 | (0.313, 0.739) | Poor [A,G] | 0.494 | (0.295, 0.693) |
| | Average [P,-] | -0.102 | (-0.268, 0.064) | Average [P,-] | -0.096 | (-0.249, 0.058) |
| | Good [P,-] | -0.432 | (-0.667, -0.198) | Good [P,-] | -0.407 | (-0.624, -0.190) |
| **IFIS flexibility** | Poor [-,-] | 0.420 | (0.217, 0.624) | Poor [A,G] | 0.375 | (0.185, 0.566) |
| | Average [-,G] | -0.045 | (-0.213, 0.122) | Average [P,G] | -0.043 | (-0.199, 0.113) |
| | Good [-,A] | -0.533 | (-0.784, -0.283) | Good [P,A] | -0.470 | (-0.704, -0.236) |
| **Muscle volume (Z-score)** | | | | | | |
| **IFIS overall fitness** | Poor [-,-] | -0.052 | (-0.399, 0.295) | Poor [-,G] | -0.231 | (-0.546, 0.085) |
| | Average [-,-] | -0.115 | (-0.297, 0.066) | Average [-,G] | -0.146 | (-0.310, 0.017) |
| | Good [-,-] | 0.139 | (-0.049, 0.327) | Good [P,A] | 0.225 | (0.054, 0.395) |
| **IFIS cardiorespiratory fitness** | Poor [-,-] | -0.023 | (-0.199, 0.152) | Poor [-,G] | -0.130 | (-0.293, 0.032) |
| | Average [-,-] | -0.088 | (-0.290, 0.115) | Average [-,-] | 0.033 | (-0.153, 0.220) |
| | Good [-,-] | 0.286 | (-0.028, 0.600) | Good [P,-] | 0.337 | (0.052, 0.623) |
| **IFIS muscular fitness** | Poor [-,G] | -0.502 | (-0.816, -0.188) | Poor [A,G] | -0.566 | (-0.851, -0.281) |
| | Average [-,G] | -0.090 | (-0.272, 0.092) | Average [P,G] | -0.055 | (-0.220, 0.110) |
| | Good [P,A] | 0.257 | (0.076, 0.438) | Good [P,A] | 0.243 | (0.079, 0.407) |
| **IFIS speed-agility** | Poor [-,-] | -0.160 | (-0.388, 0.067) | Poor [-,G] | -0.217 | (-0.426, -0.008) |
| | Average [-,-] | -0.008 | (-0.185, 0.170) | Average [-,-] | 0.034 | (-0.128, 0.196) |
| | Good [-,-] | 0.206 | (-0.043, 0.455) | Good [P,-] | 0.192 | (-0.035, 0.418) |
| **IFIS flexibility** | Poor [-,G] | -0.184 | (-0.401, 0.033) | Poor [-,G] | -0.185 | (-0.384, 0.014) |
| | Average [-,-] | 0.007 | (-0.170, 0.183) | Average [-,-] | 0.026 | (-0.136, 0.187) |
| | Good [P,-] | 0.257 | (-0.006, 0.520) | Good [P,-] | 0.216 | (-0.026, 0.457) |

Data are presented as mean and 95% confidence intervals. Adjusted models are adjusted by age and sex. Superscripts indicate statistically significant Tukey's pairwise comparisons ($p<0.05$) for the Z-scores of body composition outcomes across categories of the IFIS scores: P (Poor), A (Average), and G (Good). For example, in IFIS overall fitness, for the unadjusted model, those rating their overall fitness as "Poor" had significant differences in the Z-score for body mass index compared to those rating their overall fitness as "Average" or "Good".
CI: confidence interval, IFIS: International Fitness Scale.
